# Consent in Canadian-Led Critical Care Research During the COVID-19 Pandemic: A Scoping Review

**DOI:** 10.1101/2024.02.01.24302151

**Authors:** Karla Krewulak, Lisa Albrecht, Saoirse Cameron, Jessica Gibson, Dori-Ann Martin, Rebecca Porteous, Margaret Sampson, Katie O’Hearn

**Affiliations:** Department of Critical Care Medicine, Cumming School of Medicine, University of Calgary, Calgary, AB, Canada; Children’s Hospital of Eastern Ontario Research Institute, 401 Smyth Road, Ottawa, ON, Canada; Children’s Hospital - London Health Sciences Centre, London, ON, Canada; Department of Pediatrics, Cumming School of Medicine, University of Calgary, Calgary, AB, Canada; Pediatric Intensive Care Unit, Alberta Children’s Hospital Research Institute, University of Calgary, Calgary, AB, Canada; Department of Critical Care Medicine, The Ottawa Hospital, Ottawa, ON, Canada

## Abstract

**Introduction:** Despite the importance of critical care research during the SARS-CoV-2 pandemic, several pandemic-related factors made the process of obtaining prior written informed consent for research infeasible. To overcome these challenges, research studies utilized alternate informed consent models suggested by available guidance.

**Objective:** To describe the consent models used in Canadian intensive care unit (ICU) and pediatric ICU (PICU) studies during the COVID-19 pandemic.

**Data Sources:** We searched MEDLINE, EMBASE, CENTRAL, clinicaltrials.gov, and medRxiv from 01-Jan-2020 to 28-Apr-2023 using Medical Subject Headings and keywords related to the setting (ICU, PICU), study design (e.g., RCT) and study region (i.e., Canada). We included Canadian-led studies that were enrolling during the SARS-CoV-2 and reported on consent. Two independent reviewers reviewed titles/abstracts and full text articles for inclusion.

**Results:** We included 13 studies from adult (n=12, 92.3%) and pediatric (n=1, 7.7%) populations. Some study authors reported that informed (n=3/13, 23.1%) or a priori (n=2/13, 15.4%) consent was obtained, without further details. Study authors also reported using written informed (n=4/13, 30.8%), deferred (n=3/13, 23.1%), verbal/waived/assent (each n=2/13, 15.4%), or that ethics approval was not necessary which means consent was not required (n=1/13, 7.7%). Five studies (n=5/13, 38.5%) used multiple consent models: a priori/deferred (n=2/5, 40%), written/verbal (n=2/5, 40%), or waived/assent (n=1/5, 20%).

**Conclusion:** This scoping review underscores the importance of transparent reporting of or modifications to trial procedures during crises, such as the COVID-19 pandemic. Improved reporting practices and exploration of alternate consent models, including electronic consent, are crucial for advancing critical care trials beyond the pandemic and preparing for future health emergencies.

## INTRODUCTION

On March 11, 2020, the World Health Organization declared the worldwide outbreak of the severe acute respiratory syndrome coronavirus 2 (SARS-CoV-2) a global pandemic. A substantial proportion of Canadians with coronavirus disease of 2019 (COVID-19) required intensive care unit (ICU) care. As a result, many ongoing research studies shifted to include COVID-19 patients, and many new studies specifically focused on COVID-19 were initiated. COVID-19 research became a global priority (1). At the same time, non-pandemic critical care research was still viewed as extremely important, as many of these studies would still be relevant for patients with COVID-19 (2,3).

Despite the importance of critical care research during the COVID-19 pandemic, several pandemic-related factors made the process of obtaining prior written informed consent for research infeasible. Canadian institutions implemented a work from home policy that prevented research staff from being onsite. Visitor restrictions prevented critically ill patients’ Surrogate Decision Maker (SDM), who most commonly provide consent for intensive care unit (ICU) studies (4), from being present at the patient’s bedside. When research staff were able to attend the ICU, infection control procedures prevented research staff from transferring the informed consent document into and out of a patient’s room.

To overcome these challenges, research studies utilized alternate informed consent models suggested by available guidance (e.g., remote, waived consent) (2,5). Some of these changes, such as more flexibility in how consent can be documented for regulated clinical trials, may be beneficial for critical care research after the pandemic is over. However, to identify the consent process changes that may facilitate critical care research in the long-term, we first need a better understanding of how Canadian adult and pediatric ICU (PICU) studies (non-pandemic and pandemic related) adjusted their consent models and procedures during the COVID-19 pandemic.

## Methods

### Protocol and registration

We conducted a scoping review of the literature to describe the consent models reported by Canadian ICU and PICU studies conducted during the COVID-19 pandemic. The protocol and objectives were established and registered a priori (Open Science Framework: https://osf.io/6kgha) before commencing the systematic literature search. Our intention was to contact corresponding authors for the standard operating procedures or information on how consent changed. However, with the publication of the CONSERVE statement (6), we decided to instead summarize what was reported in included studies. This scoping review is reported according to PRISMA-ScR (7).

### Eligibility criteria

Studies were included if they met the following criteria:

1. Study Design: Any original, published study including RCTs and quasi RCTs, observational cohort studies, survey studies, biobanking studies.
2. Outcome of Interest: Study reports any information about consent, including the consent model used (e.g., prior informed, waiver of consent, deferred), the consent rate (or information required to calculate the consent rate), and/or consent procedures.
3. Population of Interest: Study includes (P)ICU patients, or family members of (P)ICU patients.
4. Country of Origin: Study is Canadian led. The study can include sites from other countries but will be excluded unless the study is led or co-led by a Canadian investigative team (e.g., corresponding author is Canadian).
5. Language: Study is published in English or French.
6. Timeframe: The trial was actively recruiting during the COVID-19 pandemic, defined as March 11, 2020 (the date that the World Health Organization declared the worldwide outbreak of a global SARS-CoV-2 pandemic) to the date of the search.
7. Study Setting: Study occurs in an adult or pediatric ICU. A unit will be considered a PICU or ICU based on the authors’ definition.

### Information sources and search

We searched MEDLINE, EMBASE, and CENTRAL from 11-Mar-2020 to 28-Apr-2023 using the search strategy from a related study conducted by our research team (8). We performed manual searches in medRxiv (the preprint server for health sciences) for the terms “ICU,” “PICU,” “study,” “trial,” “canada” and “canadian.” Though our intention was to use MeSH terms and keywords related to COVID-19 (per the registered protocol), we removed these two groups of search terms to capture non-pandemic related studies. We also removed to study design MeSH and keywords to capture articles that did not fit into predefined study design filters.

### Selection of sources of evidence

Titles/abstracts screening, full-text screening, and data extraction were completed independently in duplicate using Covidence (www.covidence.org; Veritas Health Innovation, Melbourne, Australia). Conflicts were resolved through discussion until consensus was reached or by a third reviewer.

### Data items

Data were abstracted using Covidence (www.covidence.org; Veritas Health Innovation, Melbourne, Australia) and piloted in duplicate using five eligible studies. Data were abstracted by two independent reviewers in duplicate. Consensus and/or resolution of conflicts were performed by a third reviewer, where necessary. From each included study, we included the following: 1) Study characteristics (study design, study population, sample size, number of sites, funding source, regulatory information), 2) Enrollment information (enrollment time windows, recruitment dates), 3) Consent information (i.e., consent model used, who obtained consent, consent rate, assent), 4) Changes due to the COVID-19 pandemic.

### Critical appraisal

Given the exploratory nature of this scoping review, we did not conduct critical appraisal of individual studies (9).

### Synthesis of results

Data is summarized descriptively, which includes quantitative (i.e., frequencies, proportions) and qualitative analysis. For qualitative analysis, two coders used an iterative process to apply the framework method (10), which included the following steps: 1) familiarizing of extracted text; 2) open coding (i.e., line by line coding of extracted text); 3) developing an analytical framework; 4) applying the analytical framework to the remaining extracted text; and 5) interpreting the data.

## Results

Of 5,538 unique studies, 13 met inclusion criteria (**Figure 1**). Nearly all studies (n=12/13, 92.3%) were conducted in a critically ill adult population. This included pandemic (n=7/13, 54%) and non-pandemic (n=6/13, 46%) related studies. Most studies were randomized controlled trials (n=6/13, 46.2%) or prospective cohort studies (n=4/13, 30.8%).

**Figure 1.**
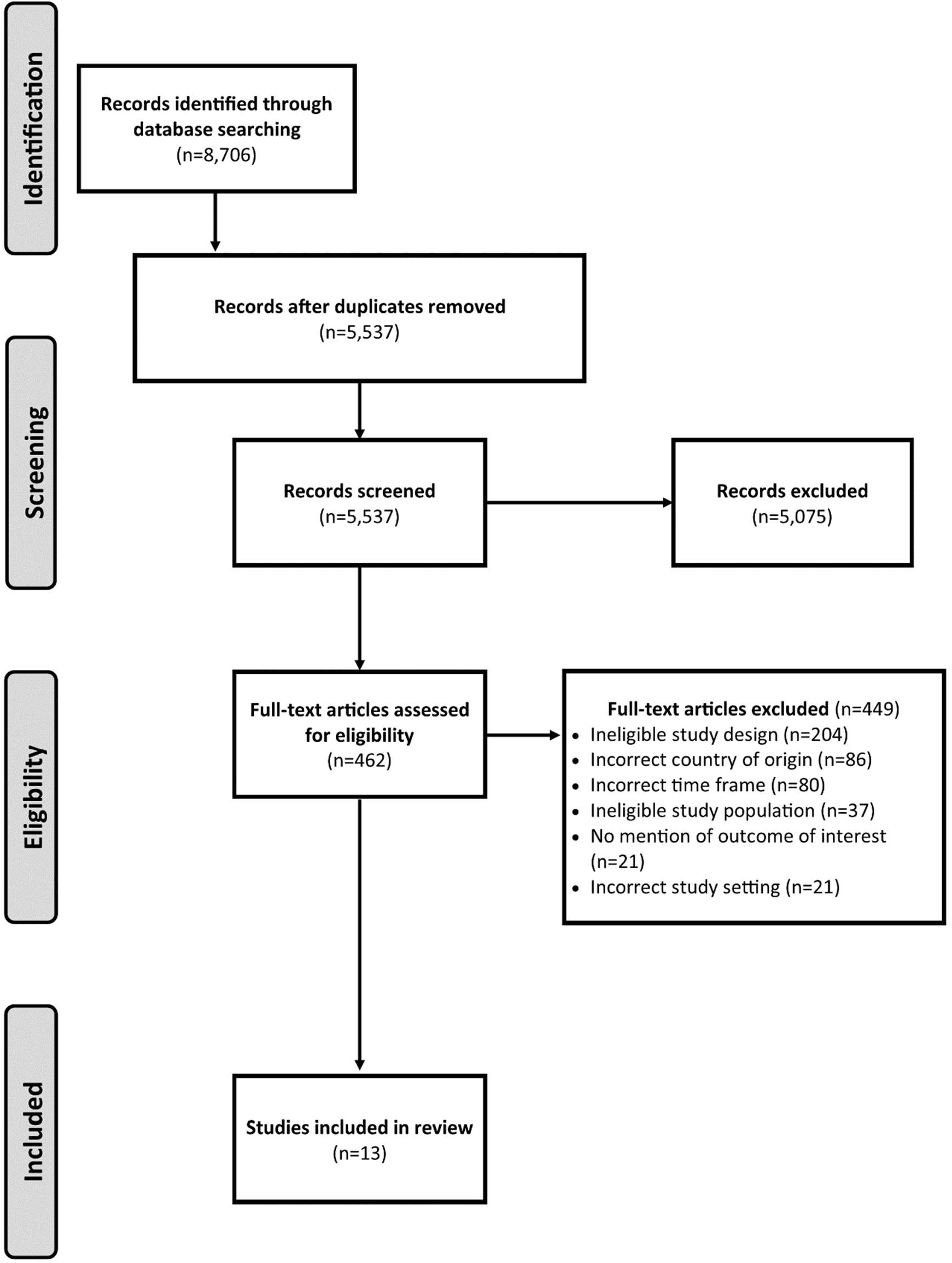
PRISMA diagram

Some studies reported that informed (n=3/13, 23.1%) or a priori (n=2/13, 15.4%) consent was obtained, without further details. Studies also reported using written informed (n=4/13, 30.8%), deferred (n=3/13, 23.1%), verbal/waived/assent (each n=2/13, 15.4%; assent from PICU youth), or that ethics approval was not necessary which means consent was not required (n=1/13, 7.7%). Five studies (n=5/13, 38.5%) used multiple consent models. This included a priori/deferred (n=2/5, 40%), written/verbal (n=2/5, 40%), or waived/assent (n=1/5, 20%). Only three (n=3/13, 23%) of the included studies began recruitment prior to the declaration of the pandemic on March 14, 2020. Though all three studies provided detail on the consent model used, none reported if the consent model was changed in response to the pandemic. As such, information on participant recruitment before/during the pandemic was also not reported. Information on included studies is displayed in **Table 1**.

**Table 1.**
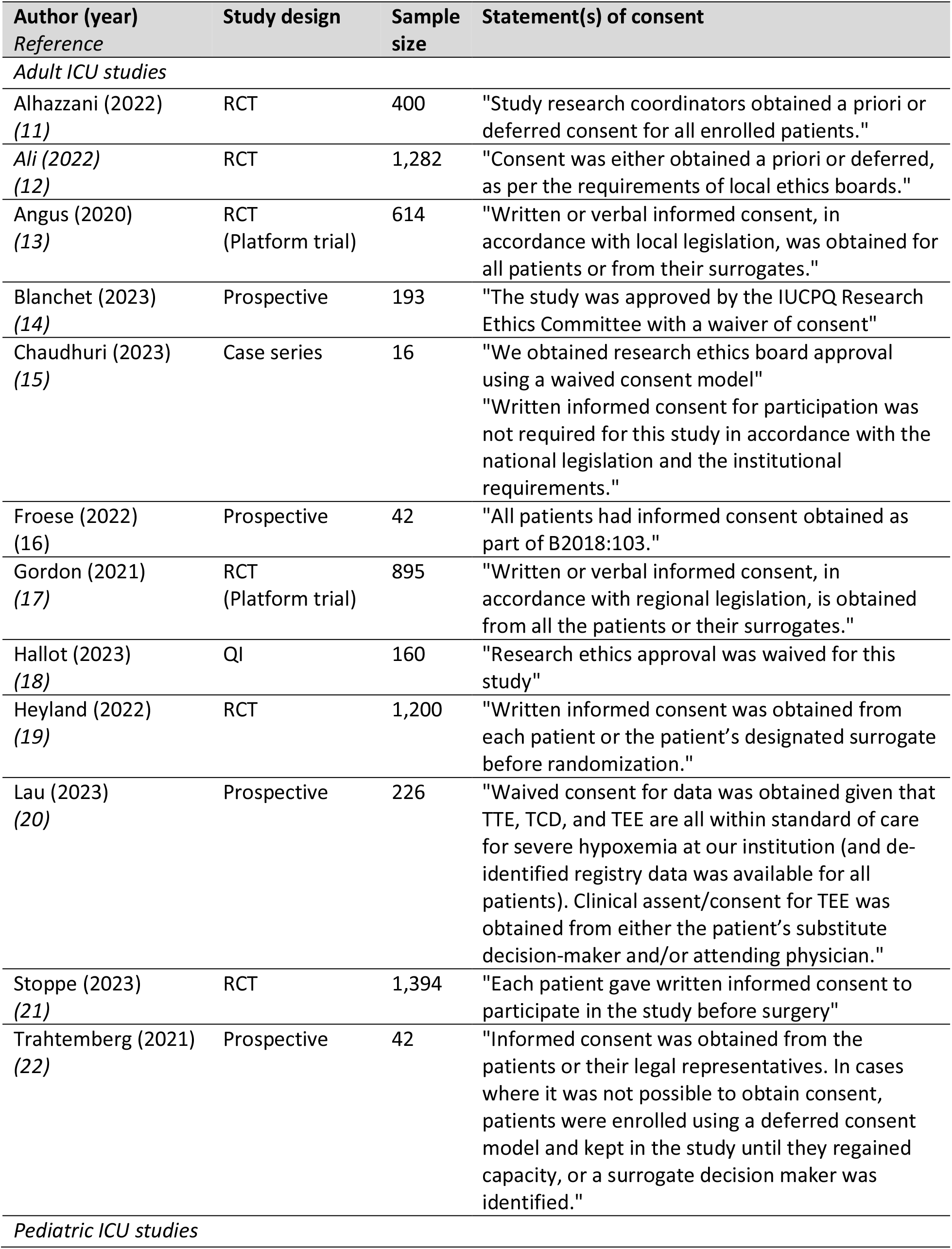
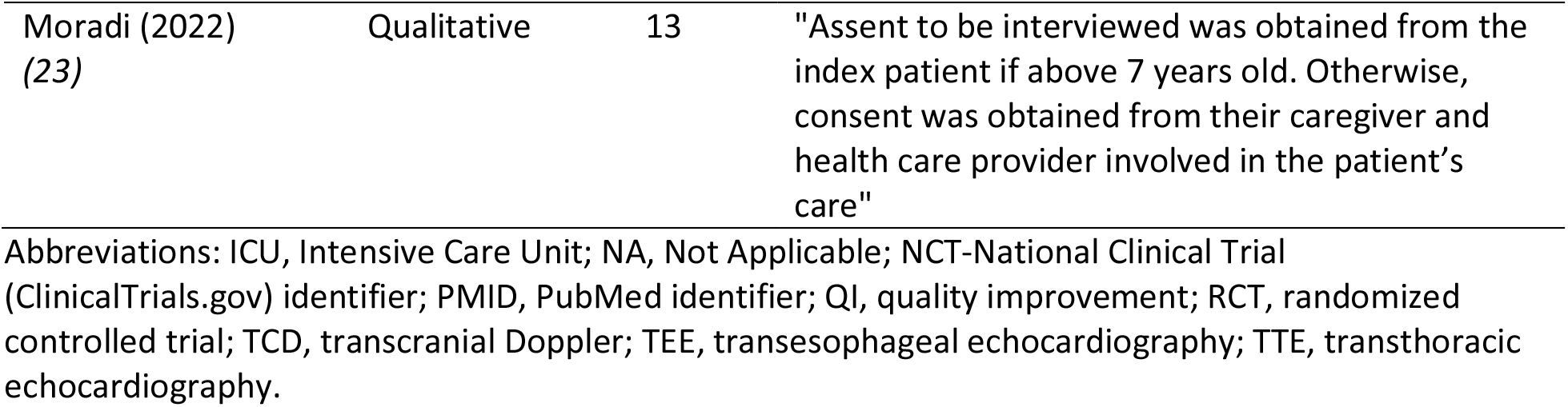
Study characteristics

Nine of the included studies (n=9/13, 69.2%; non-pandemic related: n=5; pandemic related: n=4) reported consent rate or included data to calculate consent rate. The consent rate for these nine studies ranged from 71% to 98% (non-pandemic related: 71%-98%; pandemic related: 76%-98%). The median (interquartile range [IQR]) consent rate for non-pandemic related studies was 81% (72%-81%) and for pandemic-related studies was 97% (92%-98%).

## Discussion

The COVID-19 pandemic necessitated rapid changes to consent practices in Canadian (P)ICU studies while upholding ethical standards, to ensure that patient autonomy was not compromised while following institutional pandemic guidelines around physical distancing. The current study reports on the consent models used for trials during the pandemic. Although we hoped to identify how consent processes were changed as a direct result of the pandemic, no included manuscripts reported this.

A recent study described the consent rate for an observational research study enrolling pediatric cardiac ICU patients pre-pandemic (94%) and during the one-visitor policy during the COVID-19 pandemic (79%) (25). The studies included in our current scoping review did not report changes to consent rate pre- and during COVID-19. As this is reported in future work, it will be important to understand the reason for change to consent rate (e.g., due to the stress of the pandemic or the change in consent conversations with SDMs [e.g., telephone]), and if alternate consent models (e.g., electronic consent) could mitigate these effects.

Recent guidelines emphasize the importance of/need for transparent reporting of how trial procedures are modified during extenuating circumstances like the pandemic (6). This reporting is important to understand how/which protocol modifications can benefit research longer term (after the pandemic). These changes, such as greater acceptability of remote consent and alternate consent models (24), have the potential to enhance critical care trial feasibility and accelerate recruitment (e.g., when SDMs are not present in person) beyond the pandemic.

This scoping review had strengths and limitations. The strengths include that this manuscript is reported according to PRISMA-ScR, that the search strategy was previously developed by an experienced health information specialist (MS) who is a co-author on this manuscript, and that the scoping review was conducted by and manuscript coauthored by (P)ICU research coordinators from across Canada.

Limitations include that the scoping review focuses on consent models in Canadian-led trials and cannot be generalized to other countries which may have used different consent models.

## Conclusion

This scoping review reports on the consent models employed in Canadian-led trial recruiting during the COVID-19 pandemic. Although we hoped to identify how consent processes were changed as a direct result of the pandemic, no included studies reported this. As future work unfolds, understanding the changes to consent models, such as electronic consent, is important for enhancing the feasibility and efficiency of critical care trials beyond the pandemic.

## Data Availability

All data produced in the present study are available upon reasonable request to the authors

